# In-House, Rapid, and Low-Cost SARS-CoV-2 Spike Gene Sequencing Protocol to Identify Variants of Concern Using Sanger Sequencing

**DOI:** 10.1101/2021.08.09.21261723

**Authors:** Fatimah S. Alhamlan, Dana M. Bakheet, Marie F. Bohol, Madain S. Alsanea, Basma M. Alahideb, Faten M. Alhadeq, Feda A. Alsuwairi, Maha Al-Abdulkareem, Mohamed S. Asiri, Reem S. Almaghrabi, Sarah A. Altamimi, Maysoon S. Mutabagani, Sahar I. Althawadi, Ahmed A. Alqahtani

## Abstract

**Background:** The need for active genomic sequencing surveillance to rapidly identify circulating SARS-CoV-2 variants of concern (VOCs) is critical. However, increased global demand has led to a shortage of commercial SARS-CoV-2 sequencing kits, and not every country has the technological capability or the funds for high-throughput sequencing platforms. Therefore, this study aimed to develop and validate a rapid, cost-efficient genome sequencing protocol that uses supplies, equipment, and methodologic expertise available in standard molecular or diagnostic laboratories to identify circulating SARS-CoV-2 variants of concern.

**Methods:** Sets of primers flanking the SARS-CoV-2 spike gene were designed using SARS-CoV-2 genome sequences retrieved from the Global Initiative on Sharing Avian Influenza Data (GISAID) Database and synthesized in-house. Primer specificity and final sequences were verified using online prediction analyses with BLAST. The primers were validated using 282 nasopharyngeal samples collected from patients assessed as positive for SARS-CoV-2 at the diagnostic laboratory of the hospital. The patient samples were subjected to RNA extraction followed by cDNA synthesis, conventional polymerase chain reaction, and Sanger sequencing. Protocol specificity was confirmed by comparing these results with SARS-CoV-2 whole genome sequencing of the same samples.

**Results:** Sanger sequencing using the newly designed primers and next-generation whole genome sequencing of 282 patient samples indicated identical VOCs results: 123 samples contained the alpha variant (B.1.1.7); 78, beta (B.1.351), 0, gamma (P.1), and 13, delta (B.1.617.2). The remaining samples were not 100% identical to the reference genome; however, 99.97% identity indicated that there was minimal variation as the virus spread throughout the nation. Only four samples had poor sequence quality by Sanger sequencing owing to a low RNA count (Ct value >38). Therefore, mutation calls were >98% accurate.

**Conclusions:** Sanger sequencing method using in-house primers is an alternative approach that can be used in facilities with existing equipment to mitigate limitations in high throughput supplies required to identify SARS-CoV-2 variants of concern during the COVID-19 pandemic. This protocol is easily adaptable for detection of emerging variants.

## Background

In March 2020, the World Health Organization (WHO) declared a global pandemic caused by SARS-CoV-2. As of July 26, 2021, the virus had infected more than 194 million people worldwide and caused the deaths of 4.16 million individuals [1]. As of July 2021, there were four SARS-CoV-2 variants of concern and nine variants under investigation. The variants of concern were the alpha (B.1.1.7), beta (B.1.351), gamma (P.1), and delta (B.1.617.2) variants where variant alpha had been reported in 178 countries, beta in 123 countries, gamma in 75 countries, and delta in 111 countries [2].

SARS-CoV-2 is a single-stranded RNA-enveloped virus. An RNA-based metagenomic next-generation sequencing (NGS) approach was applied to characterize its entire genome, which is 29,881□base pairs in length (GenBank No. MN908947), encoding 9860 amino acids [3]. The spike (S) gene accounts for 1273 amino acids and consists of a signal peptide (amino acids 1–13) located at the N-terminus, an S1 subunit (14–685 residues), and an S2 subunit (686–1273 residues); the last two regions are responsible for receptor binding and membrane fusion, respectively [4]. Thus, the spike gene plays an important role in the attachment of the viral particle to the receptor cells in the host, range determination, and membrane fusion [5, 6]. We focused on the spike gene for this study because of its many implications in disease transmissibility, infectiousness, and immune escape as well as for its potential as a therapeutic or diagnostic target.

Genomic sequencing is the primary tool for revealing the virus blueprint and genetic code. Given the automation and commercialization of high-throughput DNA sequencing, conducting whole genome sequencing using NGS technology would be ideal under normal situations with sufficient supplies. However, owing to low supplies during the COVID-19 pandemic and a lack of expertise in this technology in some countries, scientists worldwide have turned to the gold standard in genomic sequencing: Sanger sequencing [7]. Sanger sequencing (with polymerase chain reaction [PCR]) can be conducted using equipment that typically exists in most standard molecular or diagnostic laboratories worldwide. We therefore aimed in this work to provide a rapid and cost-efficient approach to detect circulating variants of concern (VOCs) using a protocol that can be applied in any standard molecular or diagnostic laboratory. The present study successfully designed primers in-house targeting the SARS-CoV-2 spike gene and developed and validated a rapid, cost-efficient protocol for using these primers and conventional PCR combined with Sanger genomic sequencing to timely report the circulating SARS-CoV-2 variants.

## Methods

### Design and Synthesis of Oligonucleotide Primers

More than 200 complete SARS-CoV-2 genome sequences were retrieved from the Global Initiative on Sharing Avian Influenza Data (GISAID) database (https://www.epicov.org/epi3/cfrontend#lightbox-814829872) December 2020 and were aligned using the Clustal W algorithm of the MegAlign module to identify the conserved regions using DNAStar software (DNASTAR; Madison, WI). Oligonucleotide primers were designed for the SARS-CoV-2 spike genes to ensure maximal efficiency and sensitivity. The desired primers were designed using the consensus sequences from different SARS-CoV-2 isolates from around the world, and the genome sequence from the first virus detected in Wuhan was used as a reference (accession No. MN908947). Primer3 online tools (version 4.1.0) (https://primer3.ut.ee/) was used for primer design. Because a successful PCR assay requires efficient and specific amplification, the primers were assessed for several properties, including melting temperature, secondary structure, and complementarity. Primers required a GC content of 50%-60%, a melting temperature between 50 °C and 65 °C, a salt concentration of 50 mM, and an oligonucleotide concentration of 300 nM. The specificity of the primers and final sequences were verified using in silico prediction analyses with the online Basic Local Alignment Search Tool (BLAST) (https://blast.ncbi.nlm.nih.gov/Blast.cgi). None of our designed primers showed genomic cross-reactivity with other viruses, the human genome, or other probable interfering genomes in the BLAST database analysis [8]. Primers were synthesized in-house at the Oligonucleotide Synthesis Unit of the Center for Genomic Medicine at King Faisal Specialist Hospital and Research Centre.

### Sample Collection and Ethical Considerations

This study was performed in compliance with all applicable national and international ethical guidelines for conducting research on human participants, including in accordance with the Code of Ethics of the World Medical Association (Declaration of Helsinki), and was approved by the institutional review board at King Faisal Specialist Hospital and Research Centre (KFSHRC) (IRB #220 0021). This board also granted a waiver for obtaining informed consent owing to the use of deidentified samples for this study.

In total, deidentified 282 nasopharyngeal swabs were collected from the CAP-accredited diagnostic laboratory in the Pathology and Laboratory Medicine Department at KFSHRC. These samples were confirmed positive for the presence of SARS-CoV-2 with Ct values ranging from 14 to 40.

### Nucleic Acid Extraction and cDNA Synthesis

In total, 282 samples obtained by nasopharyngeal swab and collected in universal viral transport medium were used in the nucleic acid extraction with a MagMAX Viral/Pathogen Nucleic Acid Isolation Kit (catalog No. A42352, Thermo Fisher Scientific) using a KingFisher Flex system (catalog No. 5400610, Thermo Fisher Scientific). Because cDNA synthesis is required to prepare patient samples for subsequent PCR, SuperScript IV VILO Master Mix (catalog No. 11766050, Thermo Fisher Scientific) was used. Briefly, a mixture of 8 μL of RNA extract (approximately 50 ng/μL), 4 μL of Master Mix, and 8 μL of nuclease-free water was incubated at 25 °C for 10 min. The RNA was reverse transcribed by incubating the mixture at 50 °C for 10 min, and the reaction was terminated at 85 °C for 5 min, and then place on ice.

The quality and the quantity of the RNA and cDNA samples were determined using a NanoDrop spectrophotometer. The ratio of sample absorbance at wavelengths of 260 and 280 nm was obtained to assess the nucleic acid purity (approximately 2.0 for RNA and approximately 1.8 for cDNA).

### PCR and Sanger Sequencing

The primer sets used are given in **Table 1**. For a total reaction volume of 25 μL, 12.5 μL of 2× GoTaq Master Mix (Promega; Madison, WI, USA), 2 μL of cDNA, 0.25 μL of sense primer (20 μM), 0.25 μL of anti-sense primer (20 μM), and 10 μL of nuclease-free water were used. The PCR cycle was conducted using a Veriti 96-Well Fast Thermal Cycler (catalog No. 4375305, Thermo Fisher Scientific) as follows: 95 °C for 5 min; then 45 cycles of 95 °C for 30 s, 58° C for 30 s, and 72 °C for 30 s; followed by 72 °C for 5 min. The positive control was a sample containing SARS-CoV-2, and the negative control was water.

**Table 1.**
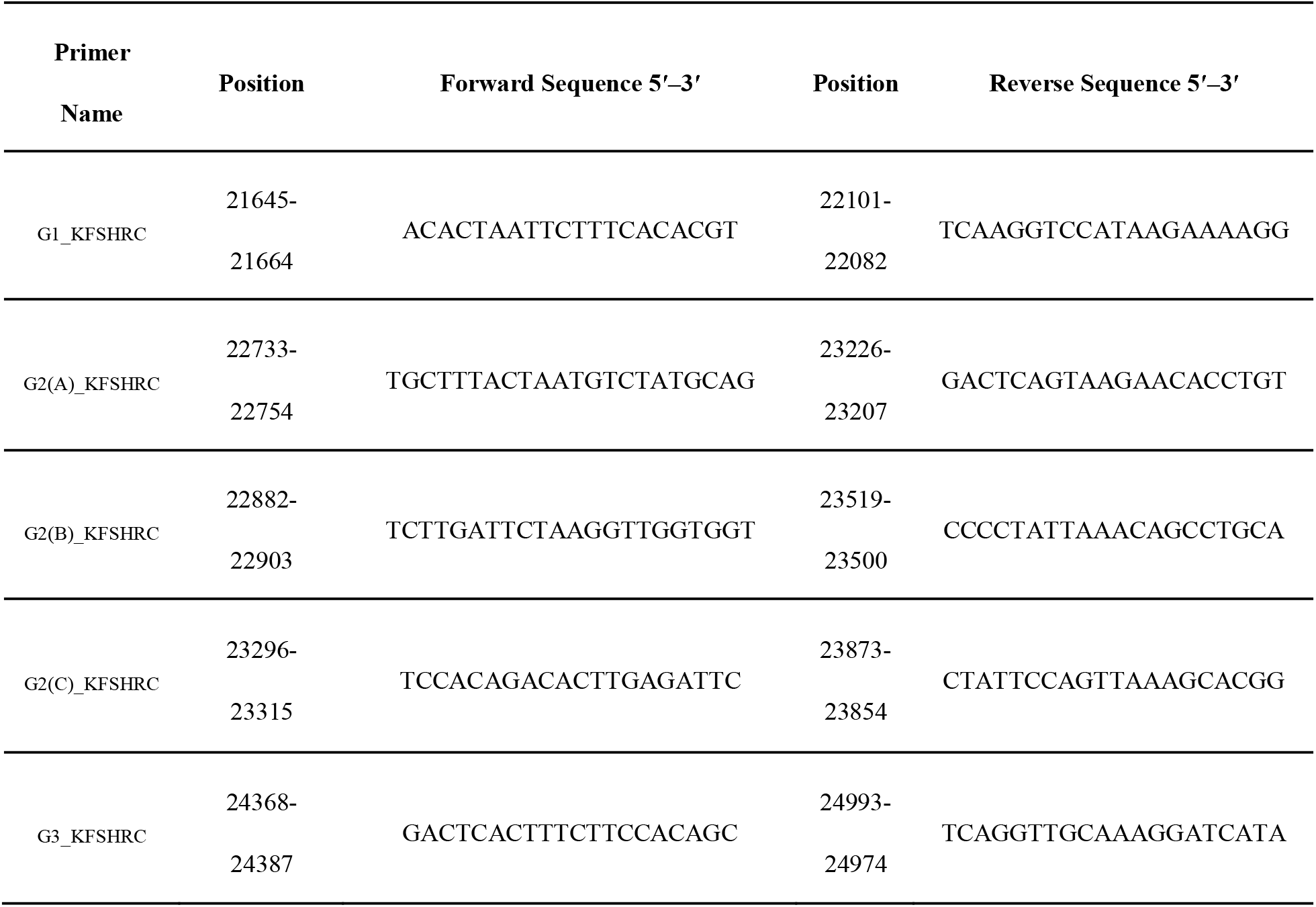
PCR primer sets for detection of SARS-CoV-2 variants of concern

The PCR products were separated using 1.5% agarose gel electrophoresis and visualized using ultraviolet light (Gel Doc EZ System; Bio-Rad). The retained PCR products were purified using AMpure XP and then sent for Sanger sequencing to the Core Facility of the Center for Genomic Medicine at KFSHRC. For Sanger sequencing, sequence chromatograms of 282 spike gene sequences were aligned using the Lasergene suite for sequence analysis (DNASTAR), with the WH-human1 SARS-CoV-2 sequence (MN908947) used as a reference [9].

### SARS-CoV-2 Whole Genome Sequencing

Portions of the same 282 nasopharyngeal samples used for nucleic acid extraction and cDNA synthesis were also subjected to nucleic acid extraction using a MagMAX™ Viral/Pathogen Nucleic Acid Isolation Kit (Cat No. A42352, Thermo Fisher Scientific; MA, USA) for whole genome sequencing using the Ion AmpliSeq SARS-CoV-2 Research Panel with the Ion Torrent S5 platform by following the manufacturer’s instructions (Thermo Fisher Scientific). For SARS-CoV-2 whole genome sequencing, variants were detected using the S5 Torrent Suite™ software Variant Caller (TVC), version 5.12.

## Results

Our primers and protocol were validated by first determining that the bands with the corresponding amplicon size were visualized for positive control samples, whereas for negative samples, the band was absent. Of samples from 282 patients, 282 samples showed the correct band of the corresponding size, however four samples with high Ct values (≥38) showed faint bands. These results indicated that our primers were specific and provided positive results with samples that were previously confirmed being positive. Our results were 100% concordant with those of the diagnostic laboratory at KFSHRC, which used the DiaPlexQ™ Novel Coronavirus (2019-nCoV) Detection Kit. The PCR product of 282 samples, including the samples with faint bands, were sent to the sequencing core facility to perform Sanger sequencing.

The output sequences of 282 patient samples were analyzed using DNAStar Lasergene Suite. Multiple sequence alignment of the spike gene was conducted with the reference genome (GenBank Protein Accession: QHD43416.1). The sequencing results were as follows: 123 samples contained the alpha variant, 78 samples contained the beta variant, no sample contained the gamma variant, 13 samples contained the delta (B.1.617.2), 64 samples were non-VOCs that belonged to none of these variants and 4 samples had poor sequence. The major mutations detected were as follows: 69-70Del, 144Del, N501Y, A570D, P681H, T716I, S982A and D1118H for the alpha variant; D80A, K417N, E484K, N501Y and A701V for the beta variant; and G142D, 156-157Del, R158G, L452R, T478K, P681R, and D950N for the delta variant (Figure 1). Moreover, the remaining samples were non-VOCs that belonged to none of these variants and had 99.97% identity with the reference genome. These mutations are not currently considered variants of concern nor variants under investigation. Of note, the D614G mutation was the most predominant in this cohort. Using the generated primers and developed protocol, we were able to sequence >98% of the positive samples with accurate mutation calls that matched the NGS data.

**Figure 1.**
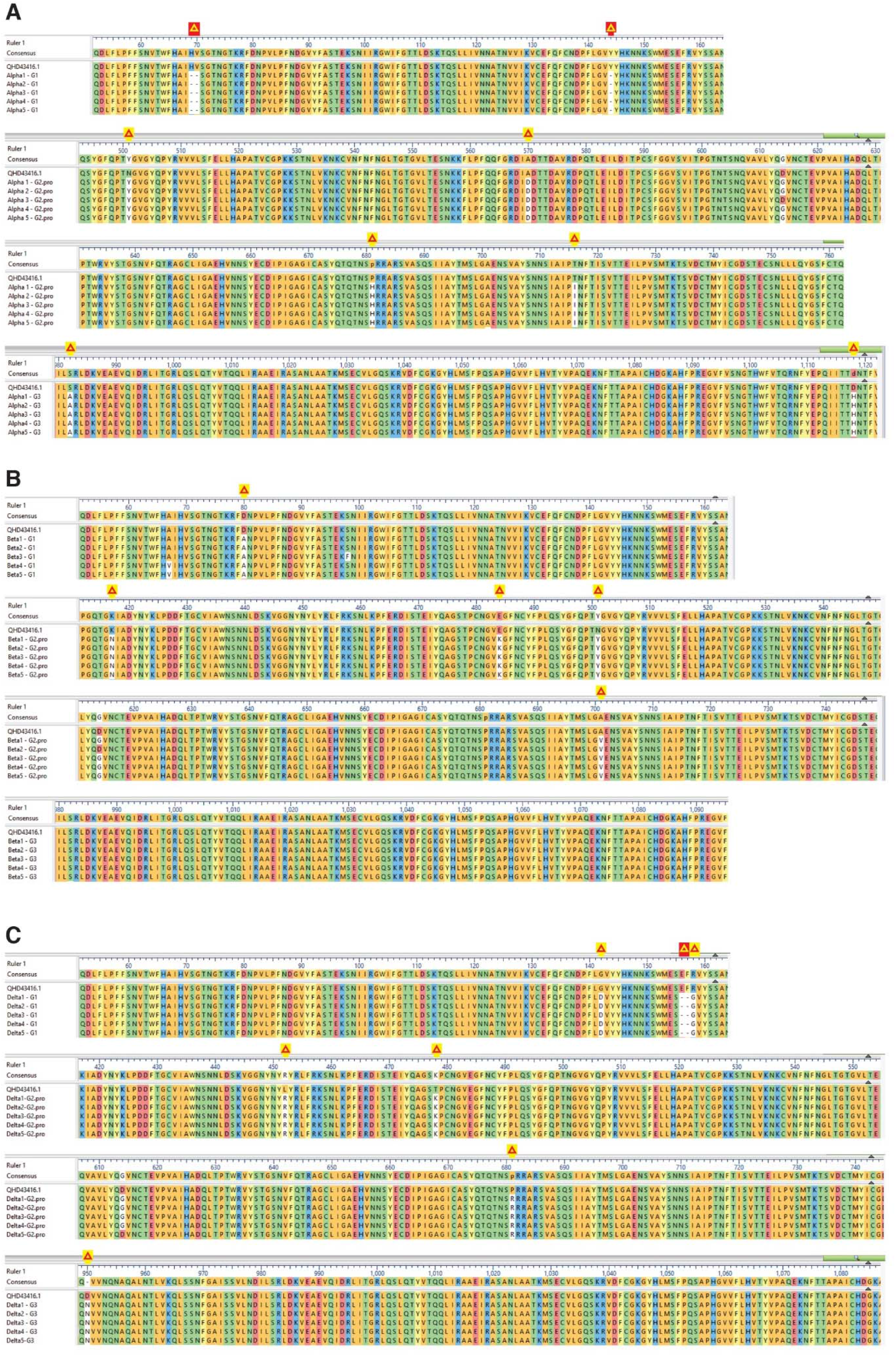
Aligned amino acid residues of the spike protein of SARS-CoV-2 from representative patient samples against the reference genome. Yellow triangles indicate deletions while red triangles indicate major mutations at different positions. **A:** represents alpha variant with major mutations such as 69-70Del, 144Del, N501Y, A570D, P681H, T716I, S982A, D1118H. **B:** represents beta variant with major mutations such as D80A, K417N, E484K, N501Y, A701V. **C:** represents delta with major mutations such as G142D, 156 -157 Del, R158G, L452R, T478K, P681R and D950N. Reference sequence: GenBank Protein Accession: QHD43416.1. [SARS-CoV-2 isolate Wuhan-Hu-1, complete genome]

To test the accuracy of and to validate the PCR and Sanger sequencing results, parallel tests to detect SARS-CoV-2 were conducted with whole genome sequencing using the Ion AmpliSeq SARS-CoV-2 Research Panel with the Ion Torrent S5 platform. For NGS data analysis, S5 Torrent Server VM was used to check the quality parameters, summary, and unaligned reads with high ISP density. Our samples showed over 70% IPS density. To check the coverage analysis, mapped reads were around 500,000 reads which indicates over 3000 coverage (Mean Depth). Our 282 samples reported ≥96% On Target Scores. Consistent with the results of the in-house primers and Sanger sequencing assay, whole genome sequencing indicated that of 278 samples, 123 samples contained the alpha variant, 78 samples contained the beta variant, no samples contained the gamma variant, and 13 samples contained the delta. For the four samples that failed to be sequenced by the Sanger approach, one sample contained the alpha variant, and three samples contained a few mutations that were not of concern.

## Discussion

Given the global decreased availability of commercial SARS-CoV-2 sequencing kits, we designed, synthesized, and validated SARS-CoV-2 spike gene primer sets to identify current variants of concern in an efficient, cost-effective, and timely manner. We specifically designed and validated these primer sets to make them available for use in other laboratories worldwide. This is a low-cost, easily performed protocol, and the reagents, laboratory supplies, and equipment are currently available as well as already found in most pathology or diagnostic laboratories.

The COVID-19 pandemic has underscored the importance of genomic surveillance as a valuable tool that quickly and effectively informs governments regarding public health decisions. However, these surveillance efforts are scattered globally owing to limited supplies and lack of sequencing infrastructure and trained personnel, especially in low- and middle-income countries [10]. Indeed, a lack of accurate and fast SARS-CoV-2 genomic results may prolong the pandemic. According to the U. S. Centers for Disease Control and Prevention, genomic sequencing is an important approach that allows scientists to identify SARS-CoV-2 and its variants and monitor how they change over time into new variants, to understand how these changes affect the characteristics of the virus, and to use this information to better understand how it might impact public health [11]. According to GISAID, most countries are falling short on the global repository of SARS-CoV-2 genomic data. Of 180 million confirmed COVID-10 cases, approximately 2 million have been submitted [9]. Thus, our sequencing protocol will help overcome the challenges of limited supplies and provide in-house sequencing primers and protocol that can be performed in many laboratories.

Our whole genome sequencing results showed high concordance with the PCR/Sanger sequencing results. Both platforms were successful in detecting the alpha, beta, and delta variants of concern. Recent studies have explored the possibility of using in-house primers and Sanger sequencing to identify SARS-CoV-2 variants. Testing with PCR has been considered the gold standard for viral gene detection, and it is widely used because of the ease in assay design and its relatively low setup and running costs. During the pandemic, several other laboratories have leveraged PCR technology for detection or sequencing to diagnose COVID-19, with infected individuals identified by the successful amplification of the viral genome obtained using nasopharyngeal swabs. Some facilities, especially those in resource-constrained settings, may lack NGS equipment, but most facilities have Sanger technology. Indeed, Shaibu at al. in Nigeria sequenced the whole SARS-CoV-2 genome using Sanger sequencing technology [12]. When researchers in China mapped the genome of the coronavirus isolated from one of the first patients in the Wuhan outbreak in December 2019 and made it available to scientists worldwide, they enabled the global development of diagnostic tests, vaccines, and drugs. Using this information, we previously developed in-house detection assay that was approved for emergency use by the Saudi Food and Drug Authority [8].

In addition, when Public Health England announced in December 2020 that their national testing laboratories using a TaqPath detection assay were showing marked increases in spike gene target failures as of late November 2020 [13], we went back to analyze our previous TaqPath assay results that were conducted from June to August 2020 at KFSHRC using the same TaqPath assay. Out of 20,000 tests, we never found negative spike gene results (data not shown). This finding provides supporting evidence for the absence of Alpha variant in our community during that time. Since then, we established our epidemiology and genomic surveillance system in our hospital to monitor the emerging SARS-CoV-2 variants.

A major strength of this study was that by focusing on detecting only the spike gene, the turnaround time was faster than that for a whole genome sequencing platform, with less than 24 hours needed to provide the results for 94 samples. However, major limitations of using conventional PCR and Sanger sequencing are that this approach is not high throughput and only sequence short reads (≤1000 bp) compared with NGS.

## Conclusions

In conclusion, the ongoing COVID-19 pandemic poses one of the greatest global threats in modern history and has triggered severe human, social, and economic costs. Efficient, rapid, and cost-effective sequencing protocols will aid in genomic surveillance. Thus, we successfully developed such as assay for detecting SARS-CoV-2 variants of concerns that can be used by laboratories that cannot afford or cannot obtain commercial high throughput sequencing kits. We found ≥98% identity between the full genome sequence generated by NGS and by our developed Sanger sequencing platform. This in-house assay offers a viable alternative approach to increase sequencing capacity. KFSHRC will provide these primers free of charge to research laboratories in Saudi Arabia. Moreover, the developed method may be beneficial beyond the COVID-19 pandemic because it can be easily adapted for use with any emerging microbe.

## Data Availability

Raw data generated in this study and primers are available on request.

## Declarations

Authors have nothing to declare.

## Ethics approval and consent to participate

This study was approved by Research Advisory Council (RAC) of King Faisal Specialist Hospital and Research Centre (RAC No. 2200021).

## Consent for publication

Not applicable.

## Availability of data and materials

Raw data generated in this study and primers are available on request.

## Competing interests

The authors declare that they have no competing interests.

## Funding

This study was funded by the KFSHRC COVID-19 grant fund (RAC No. 2200021). The funder had no role in the design of the study and collection, analysis, and interpretation of data and in writing the manuscript.

## Authors’ contributions

F-Alhamlan supervised the project and wrote the manuscript. DB led the oligonucleotide synthesis work, MB, M-Alsanea, M-Asiri ran the in-house protocol and Sanger sequencing. BA, F-Alhadeq, F-Alsuwairi, and M-Al-Abdulkareem conducted the whole genome sequencing and data analysis. RA, S-Altamimi, MM and S-Althawadi provided the positive patient samples, ran the same samples at the diagnostic laboratory, and validated our assay. AA is the principal investigator and developed the study design. All authors have read and approved the final draft of the article and have approved its submission for publication.

## Acknowledgements

We would like to extend our gratitude to the Sequencing Core Facility, Center for Genomic Medicine, King Faisal Specialist Hospital and Research Center. The support of the Research Center administration at King Faisal Specialist Hospital & Research Center is highly appreciated. We thank the Integrated Gulf BioSystems LLC team led by Shiva, Zaem, Balavenkatesh Mani and Udayaraja GK who helped with the NGS experiments.

## Notes

### Competing Interest Statement

The authors have declared no competing interest.

## References

1. WHO, Coronavirus Disease (COVID-19) Pandemic. 2020.

2. WHO, Weekly epidemiological update on COVID-19 - 13 July 2021. 2021.

3. Chan, J.F., et al., Genomic characterization of the 2019 novel human-pathogenic coronavirus isolated from a patient with atypical pneumonia after visiting Wuhan. Emerg Microbes Infect, 2020. 9(1): p. 221–236.

4. Huang, Y., et al., Structural and functional properties of SARS-CoV-2 spike protein: potential antivirus drug development for COVID-19. Acta Pharmacol Sin, 2020. 41(9): p. 1141–1149.

5. Heald-Sargent, T. and T. Gallagher, Ready, set, fuse! The coronavirus spike protein and acquisition of fusion competence. Viruses, 2012. 4(4): p. 557–80.

6. Qiang, X.L., et al., Using the spike protein feature to predict infection risk and monitor the evolutionary dynamic of coronavirus. Infect Dis Poverty, 2020. 9(1): p. 33.

7. Slatko, B.E., A.F. Gardner, and F.M. Ausubel, Overview of Next-Generation Sequencing Technologies. Curr Protoc Mol Biol, 2018. 122(1): p. e59.

8. Alhamlan FA, A.-Q.A., Bakheet DM, Bohol MF, Althawadi SI, Mutabagani MS, Almaghrabi RS, Obeid D, Development and Validation of an In-house, Low-Cost SARS-CoV-2 Detection Assay. Journal of Infection and Public Health, 2021.

9. GISAID, 2,385,336 sequence entries with complete collection date information shared via GISAID by July 26 2021. 2021.

10. Cyranoski, D., Alarming COVID variants show vital role of genomic surveillance. Nature, 2021. ISSN 1476-4687 (online)

11. CDC, What is Genomic Surveillance? 2021.

12. Shaibu, J.O., et al., Full length genomic sanger sequencing and phylogenetic analysis of Severe Acute Respiratory Syndrome Coronavirus 2 (SARS-CoV-2) in Nigeria. PLoS One, 2021. 16(1): p. e0243271.

13. PHE, Investigation of novel SARS-COV-2 variant. Variant of Concern 202012/01. 2020.

